# Assessing the Functional Impact of PfRh5 Genetic Diversity on Ex vivo Erythrocyte Invasion Inhibition

**DOI:** 10.1101/2020.09.30.20189175

**Authors:** Adam J. Moore, Khadidiatou Mangou, Fatoumata Diallo, Seynabou D. Sene, Mariama N. Pouye, Bacary D. Sadio, Ousmane Faye, Alassane Mbengue, Amy K. Bei

## Abstract

The PfRh5-Basigin ligand-receptor interaction is an essential step in the merozoite invasion process and represents an attractive vaccine target. To reveal genotype-phenotype associations between naturally occurring allelic variants of PfRh5 and invasion inhibition, we performed ex vivo invasion inhibition assays with monoclonal antibodies targeting basigin coupled with PfRh5 next-generation amplicon sequencing. We found dose-dependent inhibition of invasion across all isolates tested, and no statistically significant difference in invasion inhibition for any single nucleotide polymorphisms. This study demonstrates that PfRh5 remains highly conserved and functionally essential, even in a highly endemic setting, supporting continued development as a strain-transcendent malaria vaccine target.

---

Despite tremendous strides in malaria control, progress has plateaued and malaria is responsible for over 405,000 deaths and 228 million cases of disease globally each year, with the large majority of these cases and deaths being caused by the species *Plasmodium falciparum* (1). The need for a highly effective second generation malaria vaccine that induces cross-strain protection remains as important as ever. One challenge in the creation of a highly effective vaccine is the extensive genetic diversity of *P. falciparum. P. falciparum* invasion of human erythrocytes is an essential stage of the life-cycle and represents a stage amenable to targeting by both therapeutics and vaccines. Due to the essential nature of invasion for parasite survival, the merozoite form of *P. falciparum* uses an expanded repertoire of invasion ligands, which can be both variantly expressed and polymorphic, facilitating immune evasion and alternative receptor utilization (2). While most of the invasion ligands involved in tight junction formation are dispensable and variant, Plasmodium falciparum reticulocyte binding protein homologous 5 (PfRh5) has been shown to be essential (3). Antibodies targeting the PfRh5 receptor, basigin (BSG), are potently inhibitory across all laboratory adapted isolates from diverse geographic origins and a collection of short-term adapted *P. falciparum* field isolates (3) and anti-PfRh5 antibodies have also been shown to be braodly neutralizing (4). Vaccine-induced anti-PfRh5 antibodies from *Aotus* monkeys have been shown to be protective in a strain-transcendent manner in in vitro growth inhibition assays (GIA) and when naturally challenged with heterologous parasite genotypes (5, 6). Additionally, vaccine-induced antibodies in humans have been shown to sufficiently inhibit parasite growth across heterologous parasite genotypes in GIA assays (7, 8). A PfRh5 vaccine is currently in Phase I/IIa clinical trials.

While essential to erythrocyte invasion, PfRh5 is also highly conserved, with 16 nonsynonymous single nucleotide polymorphisms (SNPs) having been described in published data, and 35 in unpublished data from the Pf3k project (www.malariagen.net/pf3k; (9)).Of these, only five have been found at frequencies of 10% or more in any given population globally (10). Antibodies raised against PfRh5 from the 3D7 reference strain have been shown to inhibit invasion across genetic variants in growth inhibition assays (4). However, invasion assays with diverse field isolates are needed to discover novel SNPs and assess whether these genetic variations produce significant phenotypic impacts that significantly alter the invasion mechanism ex vivo. In the case of PfRh5, GIA assays have been shown to be a true mechanistic correlate of protection and so these assays, performed ex vivo with naturally occurring isolate populations, will be incredibly informative as to the degree of genotypic and phenotypic variation in the PfRh5-BSG pathway (11). In this study, we investigate genotype-phenotype association studies ex vivo in a highly endemic region of Senegal to determine whether specific polymorphisms can influence the inhibition of monoclonal antibodies (mAbs) targeting the PfRh5-BSG invasion pathway.

## Results

Of 17 successful invasion assays, we observed strong invasion inhibition across samples at 10ug/ml of anti-BSG antibody. Antibody inhibition was dose-dependent, and was very similar to that of 3D7 (Figure 1A). As antibody concentration decreased, there was more variability in the overall level of inhibition. For subsequent analyses and statistics, we separated samples into the lowest and highest quartiles according to inhibition at 10ug/ml (Figure 1C, D). This data implies that even in a highly endemic region of Senegal, ex vivo isolates remain highly sensitive to inhibition with monoclonal antibodies targeting the PfRh5-BSG pathway. To determine whether specific SNPs may be associated with levels of inhibition, we identified SNPs in these isolates through amplicon sequencing of PfRh5. Next Generation Sequencing (NGS) was performed on the 17 samples from the successful ex vivo assays. SNPs were found in 11/17 (64.7%) of the samples (Figure 2A), relative to 3D7 reference. Ten samples contained a single SNP, while one sample (320558) had 3 SNPs. Of those with SNPs, 9 contained the C203Y SNP, 2 contained the I407V SNP, 1 contained the K429N SNP, and 1 contained the V371I SNP. Sample 320558 contained the SNPs C303Y, I407V, and K429N (Figure 2A). A Mann-Whitney U Test found no statistically significant difference between the degree of invasion inhibition with anti-BSG mAb and the presence or absence of the C203Y SNP, for any concentration of antibody (Figure 2B). Using both linear regression and correlation coefficients, as well as Mann-Whitney U Test to compare the top and bottom quartiles at the 0.1, 1, and 10ug/ml antibody concentrations, we found that neither PMR, Parasitemia, nor number of SNPs was significantly associated with degree of inhibition (Supplementary Figure 1A-D). Thus, while this population demonstrated a high prevalence of SNPs in PfRh5, there is no phenotypic association of genotype with invasion inhibition.

**Fig. 1.**
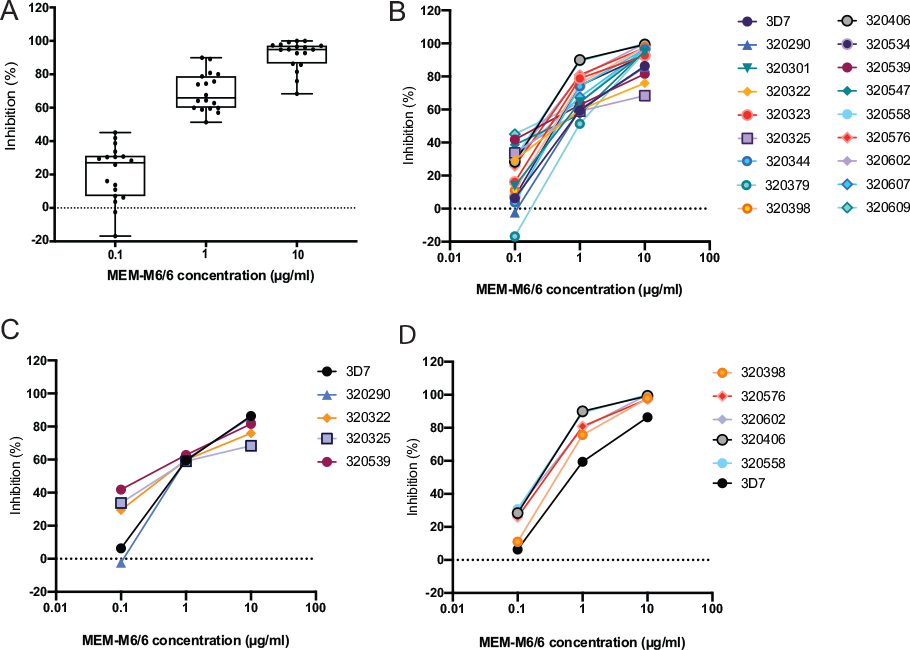
Monoclonal antibodies targeting BSG potently inhibit *ex vivo Plasmodium falciparum* isolates. Of the n=31 ex vivo invasion assays that had re-invaded and were harvested, the RPMI control wells were first counted by microscopy to determine if they reached a parasite multiplication rate (PMR) of at least 1; a quality control criteria for successful reinvasion. Seventeen of the 31 harvested assays showed a PMR of at least 1, and the inhibition data for these 17 assays are shown here. PMR is calculated as: Final parasitemia (RPMI alone)/Initial parasitemia (RPMI alone). Invasion inhibition was calculated as 100- [(Average Percent invasion in anti-BSG MEM-M6/6 antibody wells)/(Average parasitemia in IgG1 isotype control wells) * 100]. Parasitemia was counted using a Miller reticle and 4500 erythrocytes were counted for each assay. A. Distribution of inhibition at each concentration: 0.1µg/ml, 1µg/ml, and 10 µg/ml, demonstrating overall conservation in inhibition at each concentration. Box and whisker plots show the median, interquartile range, and whiskers indicate the minimum and maximum. B. Dose-response curves for inhibition with anti-BSG MEM-M6/6 antibody for each sample, including 3D7 for reference. C. Dose-response curves for inhibition with anti-BSG MEM-M6/6 antibody for samples from the bottom 25% of inhibition (calculated from the 10ug/mL concentration), including 3D7 for reference. D. Dose-response curves for inhibition with anti-BSG MEM-M6/6 antibody for samples from the top 25% of inhibition (calculated from the 10ug/mL concentration), including 3D7 for reference.

**Fig. 2.**
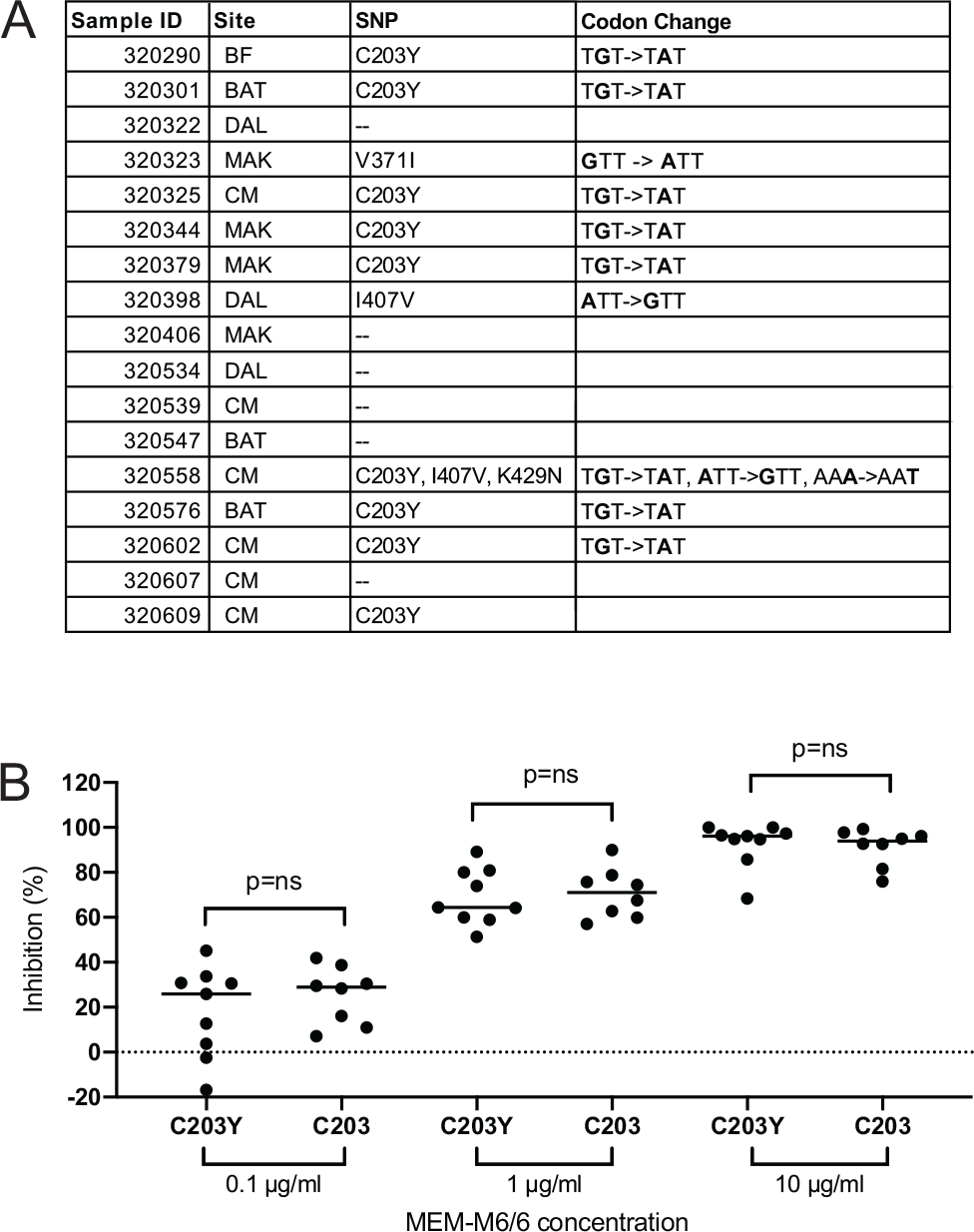
Genotype-phenotype associations identified that SNPs in PfRH5 do not influence invasion inhibition. Next Generation Sequencing (NGS) analysis of samples from successful ex vivo invasion assays show. PCR amplicons for PfRh5 exon 2 were bead-purified (Omega), quantified by Qubit and Bioanalyzer, and normalized by KAPA qPCR and bioanalyzer. Library prep was performed using unique dual indexes (UDIs), samples were multiplexed, pooled, and sequenced with a NovaSeq with targeted coverage of 500,000 reads per sample at the Yale Center for Genome Analysis. Geneious Prime Software version 2020.0.5 was used to analyze the sequencing data. Paired-end reads were trimmed with BBDuk plugin (https://www.geneious.com/plugins/bbduk; Adapter/Quality Trimming Version 38.37 by Brian Bushnell); trimmed sequences were mapped to the PfRh5 reference sequence from 3D7 (PF3D7 0424100). SNPs were called given a minimum frequency of 0.25% of all reads and coverage of minimally 5000 reads. A. Description of the single nucleotide polymorphisms (SNPs) identified in patient samples from Kedougou. Specific site of collection is abbreviated as follows: Bandafassi (BF), Bantaco (BAT), Camp Militaire (CM), Dalaba (DAL), Mako (MAK). Both nucleotide and corresponding nonsynonymous amino acid changes are indicated. Sequences are accessible in Genbank (MW042085-MW042101). Of the 11 samples with SNPs, 9 (52.9%) contained the C203Y SNP, 2 (11.8%) contained I407V, 1 (5.9%) contained K429N, 1 (5.9%) contained V371I. Sample 320558 is the only one to contain more than one SNP. Six samples contained no SNPs. These SNPs have been previously described in both published (9, 10, 12–14) and unpublished data generated by the Pf3k project (www.malariagen.net/pf3k), representing 3,248 samples from 40 separate locations in 20 countries (9). B. Genotype-phenotype associations were performed for samples with the C203Y SNP and those without (C203Y vs C203 respectively), at each concentration of anti-BSG (MEM-M6/6) antibody. Bars represent the medians of the data stratified by genotype. No statistically significant difference between either genotype was observed at any of the antibody concentrations tested.

## Discussion

This study has shown that field isolates of *P. falciparum* exhibit similar levels of invasion inhibition as lab and culture-adapted isolates when presented with varying levels of anti-BSG monoclonal antibody, supporting the hypothesis that the PfRh5-BSG invasion pathway is essential in field parasite populations and that there is minimal phenotypic variation in utilization of this pathway. Additionally, we have shown the presence of the C203Y SNP, a SNP which is present at the PfRh5-BSG interface (15), does not have a significant impact on the parasites’ dependence on the PfRh5-BSG invasion pathway, at least in associative studies. However, recent studies have found that S197Y, which is also present at the PfRh5-BSG interface, contributes to immune evasion for a neutralizing monoclonal antibodies directed against PfRh5 (7). Further studies combining genome editing to test the role of polymorphisms in an isogenic background would allow for precise investigation of whether genotypes observed in the field can alter inhibition with antibodies targeting the receptor-ligand interface. These results show PfRh5 phenotypic function and subsequent immune function are not impacted by diversity and the PfRh5-BSG invasion pathway remains essential in field samples. Previous work had shown similar results, but in lab strains and culture-adapted samples (3). Here, we add the substantial evidence from ex vivo field samples from a highly endemic site, known to contain high levels of genetic diversity, exhibiting similar behavior. By demonstrating similar behaviors in field isolates that were observed in lab isolates, we add to the growing body of evidence that PfRh5 is an important, and functionally conserved target for a malaria vaccine. This study is limited in the respect that these ex vivo assays demonstrate the behavior of parasite populations within each patient (isolates), not individual parasite clones (strains). However, polyclonal infections are common in highly endemic regions, such as Kédougou. Demonstrating that anti-BSG antibodies can significantly inhibit invasion across a genetically diverse parasite population provides a realistic idea of the potential success of a PfRh5 vaccine in patients living in endemic regions. Our findings support the potential for strain-transcendent immunity generated in response to antibodies induced by the PfRh5 vaccine as thus far we have yet to identify any genotypes that escape potent neutralizing antibodies.

## Materials and Methods

### Study Sites

This study was conducted with ethical approval from the National Ethics Committee of Senegal (CNERS) and the Institutional Review Board of the Yale School of Public Health. Samples used in this study were collected as part of ongoing surveillance conducted by Institut Pasteur de Dakar investigating causes of febrile illness. Patients were recruited from five clinics in Kédougou, Senegal. Eligibility criteria for the main study was the presence of a fever (temperature greater than or equal to 38 degrees C) and/or a fever in the past 24 hours. If a patient tested positive for *P. falciparum* on a *Pf*-specifc HRP2/3 rapid diagnostic test (RDT), a study clinical staffer assessed eligibility and obtained informed consent. A venous blood sample of 5ml in a EDTA vacutainer was obtained from consenting, enrolled patients and transported at room temperature from the clinic to the field lab for processing; no more than 6 hours between draw and processing. Thin and thick blood smears were made for each sample to confirm mono-infection with *P. falciparum* by microscopy. Patient demographics and clinical parameters are described in Supplemental Table 1.

### *Ex vivo* invasion assays

Initial parasitemia was calculated by thin smear microscopy for each sample. Samples were washed twice with unsupplemented RPMI media and resuspended in supplemented media. All cultures were plated with a starting parasitemia between 0.25% - 1.0%, at 4% hematocrit with supplemented RPMI media. Samples with parasitemia greater than 1% were diluted to 1% parasitemia with uninfected O+ erythrocytes. Three concentrations of monoclonal anti-BSG MEM-M6/6 antibody (10g/ml, 1g/ml, 0.1g/ml) were plated for each strain, alongside IgG1 isotype control, in duplicate. Additionally, wells were plated to allow for microscopic evaluation of assay progression and morphologic staging. Assay plates were cultured in a candle jar culture in a 37°C incubator. Assays were harvested when the smear-only control wells showed that at least 95% rings (no more than 5% schizonts remained) for 100 parasitized cells counted. If this criterion was not met, the culture was placed back into the incubator and smears were periodically made until it was ready to harvest. If a sample had not progressed past the schizont stage 96 hours after starting the assay, it was discarded as it did not meet the criteria for a successful assay. Upon successful reinvasion, microscopy slides were made, and counting was performed blinded. The parasitemia for the duplicate cultures were averaged and these parasitemias were used to determine parasite multiplication rate (PMR), and invasion inhibition, relative to isotype controls.

### DNA Extraction, Amplification, and Sequencing

DNA was extracted from dried blood spots (DBS) using QIAmp DNA Blood Mini Kit according to manufacturer’s instructions. PfRh5 exon 2 was PCR amplified using previously described primers (13) and high fidelity polymerase. Size and purity of the PCR amplicon was confirmed on an agarose gel prior to Next Generation Sequencing (NGS). SNPs were called given a minimum frequency of 0.25% of all reads and coverage of minimally 5000 reads.

### Statistical Analysis

GraphPad Prism 8.2.4 was used for all statistical analysis. A Mann-Whitney U Test was performed to determine if mean inhibition was significantly different for isolates with mutation or without. Statistical associations between antibody-mediated invasion inhibition and PMR, parasitemia or number of SNPs were assessed using a linear regression model and Spearman nonparametric rank correlation coefficients (data not shown).

## Data Availability

All data is presented in the paper and is freely available to the public. Sequences have been deposited in Genbank with the following range of identifiers: MW042085-MW042101

## ACKNOWLEDGMENTS

We would like to thank Lt. Dr Chares Latyr Ndiaye and Lamine Kane from Camp Militaire, Moctar Mansaly and Gerald Keita from Bandafassi, Safietou Sane and Astou Ndiaye from Mako, Adama Gueye from Bantaco, Souleymane Ngom from Dalaba, and all the healthcare workers at these sites for their partnership with Institut Pasteur Dakar. We would also like to thank the people of Kédougou for their invaluable contributions to this work. We wish to thank Laty Gaye Thiam for useful discussions and contributions to the manuscript. This publication uses data generated by the Pf3k project (www.malariagen.net/pf3k), representing 3,248 samples from 40 separate locations in 20 countries, and data published by Manske and colleagues(9).

