## Supplemental Data for "Assessing the Functional Impact of PfRh5 Genetic Diversity on Ex vivo Erythrocyte Invasion Inhibition"

**This PDF file includes:**

Fig. S1
Table S1

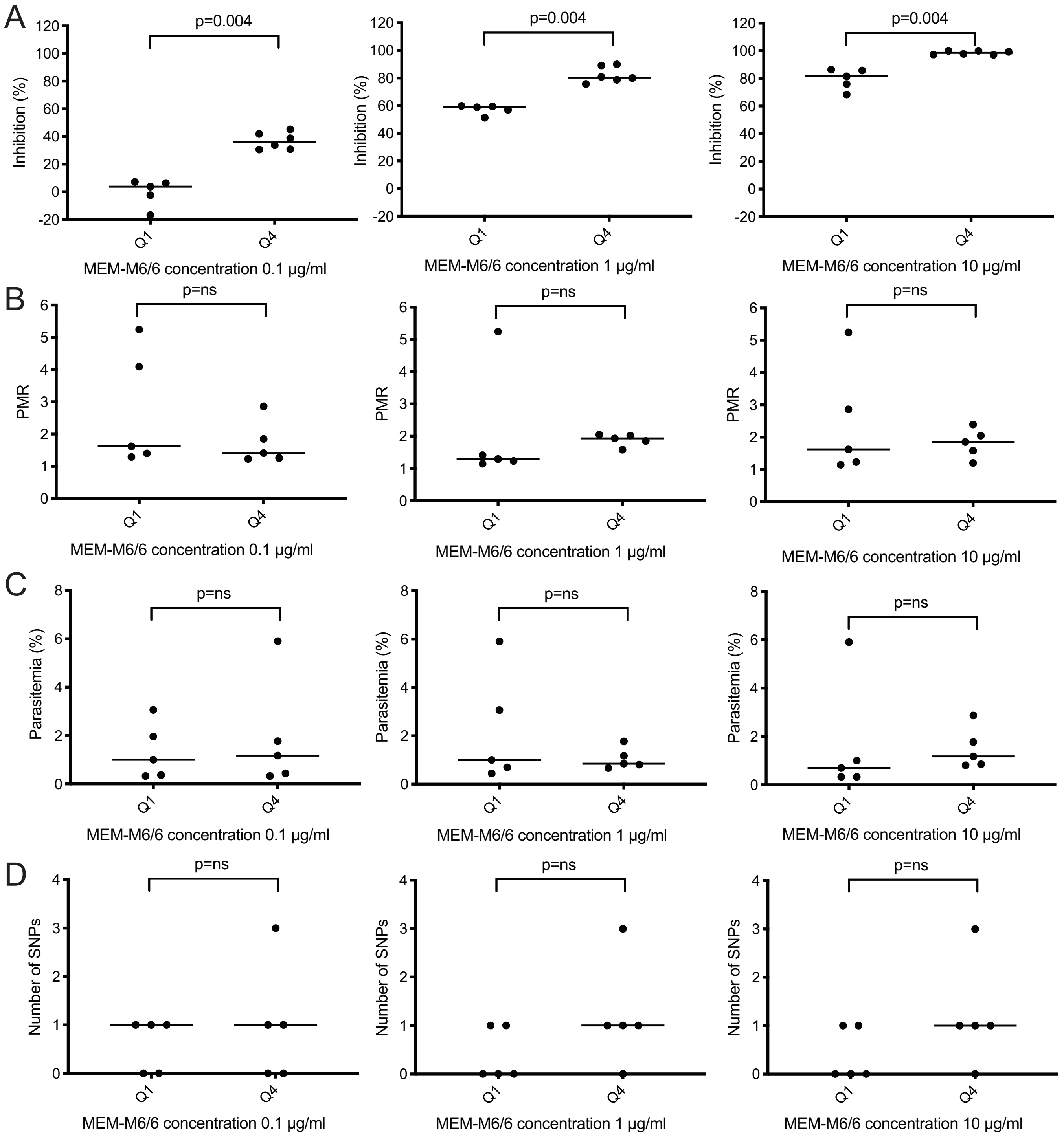


**Fig. S1. PMR, Initial Parasitemia, and Number of SNPs do not influence inhibition with mAbs to BSG.** Samples in Quartiles 1 (top 25) and 4 (bottom 25) of each antibody concentration level showed no statistically significant differences when compared by (A) Percent Inhibition (B) Parasite Multiplication Rate (PMR) (C) Percent Parasitemia (D) Number of Single Nucleotide Polymorphisms (SNPs) Bars represent the medians stratified by inhibition quartile.

| Sample | Site | Age Range, years | Sex | Temperature,ºC | Initial Parasitemia $^{a}$(%) | Adjusted Parasitemia$^{b}$(%) | Re-Invasion Parasitemia $^{c}$(%) |
|---|---|---|---|---|---|---|---|
| 320290 | BF | 35-40 | F | 38 | 0.33 | 0.33 | 0.53 |
| 320301 | BAT | 10-15 | M | 36 | 3.02 | 1.00 | 1.94 |
| 320322 | DAL | | F | 38.5 | 0.70 | 0.70 | 1.15 |
| 320323 | MAK | 10-15 | F | 39.8 | 0.67 | 0.67 | 1.35 |
| 320325 | CM | 10-15 | M | 38.4 | 5.90 | 1.00 | 1.23 |
| 320344 | MAK | 5-10 | F | 39.3 | 1.96 | 1.00 | 4.09 |
| 320379 | MAK | 15-20 | M | 38.8 | 3.06 | 1.00 | 1.29 |
| 320398 | DAL | 10-15 | M | 35.7 | 2.87 | 1.00 | 1.20 |
| 320406 | MAK | 15-20 | M | 38.6 | 0.85 | 0.85 | 1.75 |
| 320534 | DAL | 35-40 | F | 39.6 | 0.37 | 0.37 | 0.52 |
| 320539 | CM | 15-20 | F | 36.2 | 0.33 | 0.33 | 0.95 |
| 320547 | BAT | 30-35 | M | 37.8 | 0.44 | 0.44 | 0.62 |
| 320558 | CM | 10-15 | F | 39.5 | 1.18 | 1.00 | 1.85 |
| 320576 | BAT | 1-5 | F | 37 | 0.81 | 0.81 | 1.93 |
| 320602 | CM | 10-15 | F | 39.9 | 1.77 | 1.00 | 1.58 |
| 320607 | CM | 15-20 | F | 38.7 | 0.54 | 0.54 | 1.75 |
| 320609 | CM | 20-25 | M | 38 | 1.77 | 1.00 | 1.26 |

**Table S1. Patient Demographics.** Demographic data from patients enrolled in this study for which genotype-phenotype associations were performed. De-dentified sample IDs (Sample), collection site (Site), Age Range (in 5 year intervals), Sex, Temperature, Initial parasitemia, Adjusted parasitemia, and Final parasitemia. Sample distribution by site was as follows: Bandafassi (BF) 1(5.9%), Bantaco (BAT) 3(17.6%), Dalaba (DAL) 3(17.6%), Mako (MAK) 4(23.5%), Camp Militaire (CM) 6 (35.3%). All patients were *P. falciparum* positive by a *Pf*-specifc HRP2/3 rapid diagnostic test (RDT) and confirmed to be infected with *P. falciparum* only, harboring no mixed-species infections by microscopy.

a - Initial parasitemia is the percentage of infected cells at enrollment, when counting 4500 total erythrocytes by Miller reticle

b - Adjusted parasitemia is the parasitemia adjusted prior to invasion assay. Samples with initial parasitemia greater than 1% were diluted to 1% with uninfected O+ erythrocytes

c - Re-invasion parasitemia is the percentage of infected cells after assay harvest in the RPMI only (no antibody) wells, when counting 4500 total erythrocytes by Miller reticle. Sextuplicates were averaged.
